# The Infectious Diseases Clinical Research Program Acute Respiratory Infection Repository Protocol: Opportunities to Understand Current and Future Epidemics

**DOI:** 10.1101/2024.11.07.24316567

**Authors:** Simon D. Pollett, Rhonda E. Colombo, Stephanie A. Richard, Tahaniyat Lalani, Brianne Barton, Allison Malloy, Anthony Fries, Edward Parmelee, Scott Merritt, Mark Fritschlanski, Edward E. Mitre, Eric D. Laing, Kathleen Pratt, Eric C. Garges, Katrin Mende, Mark Simons, Brian Agan, David Tribble, Robert O’Connell, Timothy H. Burgess

## Abstract

**Background:** Acute respiratory infections (ARI) are a major cause of morbidity and lost workdays in both military and non-military populations. To better understand these infections and their outcomes, the Infectious Diseases Clinical Research Program has enabled nine major ARI clinical research protocols in the last decade, including observational studies and trials, spanning emerging and reemerging ARI threats including SARS-CoV-2, influenza, adenovirus, entero/rhinovirus, human metapneumovirus, respiratory syncytial virus, and other pathogens. These protocols have resulted in epidemiological, clinical and laboratory data and biospecimens from over 26,000 participants, most of whom were beneficiaries of a geographically distributed Military Health System.

**Methods:** The Acute Respiratory Infection Repository Protocol establishes a unique Department of Defense (DoD) research resource through the pooling of data and specimens from nine ARI protocols into a master, standardized database with a linked specimen repository. This will enable further targeted scientific questions in participant-level pooled meta-analyses and will serve as an on-demand repository for rapid assay development, sample size estimations for prospective studies, and observational study/clinical trial design (including as part of future rapid pandemic research response). Accordingly, the objectives and study design of this protocol are broad. This protocol will allow analyses on outcomes including: (i) short-term ARI outcomes such as hospitalization, work days lost, symptom severity and duration ; (ii) post-acute ARI outcomes, including persistence of symptoms, return-to-health, post-ARI medical encounters; (iii) vaccine effectiveness for COVID-19, influenza, and adenovirus vaccines; (iv) ARI infection and vaccination elicited immune responses (humoral, T-cell, other); (v) therapeutic effectiveness of COVID-19 and influenza antivirals (acute symptoms, hospitalization, post-acute sequelae); (vi) effectiveness of non-pharmaceutical interventions (e.g., masking) against infection; (vii) prognostic and mechanistic host viral biomarkers which correlate with the above outcomes; (viii) ARI diagnostic assay validity and performance. This repository protocol is inherently broad in scope; the collation of standardized data and phenotype-linked specimens is a fundamental, primary objective.

**Discussion:** This protocol will support statistical and laboratory analyses, including activities related to rapid epidemic response such as assay development and rapid sample size calculations for clinical trials. A series of more specific scientific questions from current and future collaborators will leverage this joint database and specimen repository; these questions will target important aspects of ARI infection, transmission, outcomes, and treatment. Future protocols (and ongoing data from existing IDCRP protocols) will be added to this collaborative repository protocol.

## Background

Acute respiratory infections (ARI) are a major cause of morbidity and lost duty days in military populations and remain a research priority for the U.S. military [1]. ARI pose a considerable risk to the health of all Military Health System (MHS) beneficiaries, and impact the readiness of active duty service members (ADSM). ARI significantly military congregate populations in shipboard and fixed land-based facilities, including due to SARS-CoV-2, influenza, adenovirus, entero/rhinovirus, and other ARI viruses [1-3]. In military recruits and trainees, crowded living conditions, stress, and sustained exposure to ARI pathogens contribute to excess ARI risk [4-7]. Nearly a quarter of medical encounters among military recruit trainees were attributed to respiratory infections in both 2018 and 2019 [8, 9]. ARI remained the second-leading cause of medical encounters in recruits in 2020 [10]. The 2020 SARS-CoV-2 outbreak on the aircraft carrier U.S.S. Theodore Roosevelt further illustrated how an ARI caused disruption in congregate shipboard settings [11]. Influenza has also caused substantive attack rates in Navy shipboard settings [12].

There remain multiple knowledge gaps about the epidemiology, optimal detection, prediction, treatment, and prevention of ARI in MHS beneficiaries and active duty servicemembers addressing these knowledge gaps is beneficial to general public health. The overarching aim of this protocol is to establish a unique Department of Defense (DoD) research resource through the pooling of at least nine ARI protocols into a master, standardized database with a linked specimen repository. These ARI protocols have been executed under the auspices of the Infectious Diseases Clinical Research Program (IDCRP), a clinical research center within the Department of Preventive Medicine & Biostatistics at the Uniformed Services University of the Health Sciences (USUHS) [13].

The ARI Repository Protocol will provide a flexible platform for statistical and laboratory analyses, including activities related to rapid epidemic/pandemic response (e.g., assay development, rapid sample size calculations for clinical trials). Future IDCRP ARI protocols (and new data from ongoing IDCRP ARI protocols) will be added to this repository protocol which will remain open indefinitely. It should be noted that this repository protocol is inherently broad in scope and the collation of standardized data and phenotype-linked specimens is a fundamental, primary objective. The joint database and specimen repository can then be leveraged to answer a series of more specific scientific questions which will target important aspects of ARI infection, transmission, outcomes, treatment, and prevention to inform Force health protection and general public health.

## Methods/design

### Study Population and Eligibility Criteria

Initially, this study involves the use of existing data and specimens from nine IDCRP ARI study protocols (**Tables 1-3**). These protocols include: The Acute Respiratory Infection Consortium study (ARIC; IDCRP-045) [14, 15]; the Immunogenicity of Novel H1N1 Vaccination among HIV-Infected Compared to HIV-Uninfected Persons (IDCRP-053) study [16]; the Epidemiology, Immunology and Clinical Characteristics of Emerging Infectious Diseases with Pandemic Potential study (EPICC, IDCRP-085) [17-36]); the Pragmatic Assessment of Influenza Vaccine Effectiveness in the DoD study (PAIVED, IDCRP-120) [38]; the COVID-19 Antibody Prevalence in Military Personnel Deployed to New York study (CAMP NYC, IDCRP-125; data only) [39]; the Prospective Assessment of SARS-CoV-2 Seroconversion study (PASS, IDCRP-126) [28, 40, 41]; the Seroprevalence of novel coronavirus antibodies among personnel deployed on the USNS Comfort during the COVID-19 pandemic study (COMFORT, IDCRP-128) [42]; The Observational Seroepidemiologic Study of COVID-19 at the United States Naval Academy study (TOSCANA, IDCRP-129) [43]; and the Prospective Investigation of SARS-CoV-2 / COVID-19 Epidemiology and Serology study (PISCES, IDCRP-130).

All studies included in the repository protocol have been conducted at military treatment facilities, in military training settings, within other DoD facilities, and/or in the context of DoD deployments related to the COVID-19 response. The overall population is therefore predominantly MHS beneficiaries, including (ADSM), retirees, and/or dependents. The nine studies included in this Repository protocol have had varying eligibility criteria (**Table 2**) but generally fall under an overarching repository eligibility criteria of MHS beneficiaries, including ADSM and those in congregate settings, who are infected by, exposed to, tested for, and/or vaccinated against ARI pathogens.

### Recruitment and consent

The IDCRP Acute Respiratory Infection Repository Protocol does not recruit or enroll new subjects. Data and specimens will only be used from participants who provided consent for secondary use of data and/or specimens, and who did not withdraw from, the original studies contributing to the repository.

### Primary objective

The primary goal of the repository protocol is to create a curated, standardized dataset and biospecimen repository incorporating multiple IDCRP ARI protocols. This will permit answering further targeted scientific questions in pooled analyses and will serve as an on-demand repository for rapid assay development, sample size calculations, and observational study/clinical trial design (including those required as part of future rapid ARI pandemic research response).

### Secondary objectives (supported and facilitated through the Primary Objective)

1. Compare the acute and chronic clinical outcomes and symptom phenotypes of ARI viral pathogens of importance to servicemembers and other MHS beneficiaries, including variant to variant comparison
2. Estimate predictors of acute and chronic clinical outcomes of ARI viral pathogens, including demographics, comorbidities, other host factors, and viral factors including infecting variant
3. Estimate and predict COVID-19, influenza, and adenovirus vaccine effectiveness in servicemembers and other MHS beneficiaries, as applicable
4. Identify predictors of optimal ARI vaccine immune responses (including those elicited by COVID-19 and influenza vaccines)
5. Estimate therapeutic effectiveness for licensed COVID-19 and influenza antivirals
6. Estimate effectiveness of non-pharmaceutical interventions, including masking and restriction of movement, against infection risk
7. Identify and evaluate prognostic and mechanistic viral and host biomarkers which correlate with the above secondary objectives
8. Evaluate the validity of current and future diagnostic and biomarker assays for the detection or prognostication of ARIs, including the support of new ARI diagnostic and biomarker assay development
9. Additional objectives as urgent DoD ARI research needs arise (e.g., during an outbreak or pandemic)

### Sample size

Current sample size is up to 27,231 participants from at least nine existing IDCRP ARI Protocols (see **Table 1**). The sample size is fixed the number of enrolled and eligible participants in the protocols being pooled. The observable effect sizes will vary by endpoints of specific pooled analyses (see Secondary Objectives) that will leverage this repository protocol.

### Data Curation

The repository protocol will only contain data and specimens from participants who approved future use of data and specimens in their consent. Data will be combined into a unified database, utilizing existing and newly created variables. A Data Management Plan (DMP) and cross protocol data dictionary will be established, ensuring data and specimens are only included in those participants who indicated consent for future use (or for studies with a waiver of consent). This multi-relational database will be developed in Oracle and managed by the IDCRP Data Coordinating Center (DCC) and will include clinical, demographic, and existing laboratory data.

Following data consolidation, a statistical analysis plan (SAP) will be developed for a preliminary assessment of the repository data to evaluate metadata structure including the total number of combined variables (and whether these are continuous, binary, or categorical), total number of combined observations, total number of common data variables (and respective number of observations) across two or more protocols, and total number of variables (and respective number of observations) which are similar enough to allow ontological cross-walking between two or more protocols. The overlap of laboratory assay data between protocols will also be evaluated, with assay type, performing laboratory, lab assay units, and measurement timepoints specified. Similarly, we will evaluate the overlap of similar specimen types (e.g., sera, PBMC) and timepoints of collection between protocols.

Documentation of these newly created common variables in the cross-protocol data dictionary will include: (i) common repository variable name, (ii) data type (e.g., continuous, categorical), (iii) any variable ontology cross-walking between protocols (iv) reference to the original variable names from individual protocols. The joint repository dataset will include a variable which defines the original IDCRP study in which the participant enrolled (the study population and eligibility criteria of which are summarized in **Table 2**). This will provide context of how individual participants were selected for enrollment and therefore identify potential selection biases in cross protocol analyses. As other open protocols are updated with further specimens and data, data will also be updated in the master repository protocol.

### Quality Management

A Quality Management Plan (QMP, including a data Quality Assurance (QA)/Quality Control (QC) process) will be applied to ensure the accuracy, completeness, and acceptability of data/specimens from the contributing protocols. The QMP leverages data system checks, previously performed QM activities, and limited review of key protocol activities (Informed Consent, data migration, and regulatory compliance). QM processes are designed to evolve during the lifecycle of this repository protocol, to continually mitigate previously- and newly-identified risks.

### Specimen curation

An inventory of all specimens will be created and verified, with cross-linkage to FreezerWorks a specimen freezer inventory program (Dataworks Development, Inc; Mountlake Terrace, WA). An initial report will describe the demographic, clinical and laboratory characteristics, and specimen availability. Data and specimens will be updated in the master repository protocol as other open protocols are updated with further specimens and data.

### Approach to analyses

After the scope and overlap of the data and specimens have been assessed, the consolidated data and/or specimens will be assessed to answer specific research questions as described in the secondary objectives with individual participant data and/or specimen results. In addition, the report will be used by the analysis team and collaborative investigators to help design new specific, hypothesis-driven questions that will be answered leveraging the repository protocol pooled data.

New analysis ideas will be vetted by an analysis working group (reflected by the byline authors of this paper). This is a collaborative framework in which (i) contributing PIs have oversight on the use of specimens and data from their protocols and can be part of the joint scientific analysis enterprise, (ii) competing laboratory demands for specimens can be balanced early and often, (iii) lab and statistical analyses can be prioritized based on a consensus regarding the most impactful, specific study questions. New analysis ideas will be presented to the working group, and after consensus is reached, analyses will proceed if they are covered under the scope of the existing protocol.

The approach to individual participant-level data (IPD) pooled meta-analyses (secondary objectives) will be tailored to the specific scientific question. This will follow IPD analysis best-practices as described by Riley et al. [44] and IPD meta-analysis best practices for reporting results (further described in Riley et al [44]). Importantly, such ARI Repository IPD metanalyses will not necessarily occur across all contributing protocols. Consideration of the underlying sampled population, study design, and common variables will be incorporated into a DMP and SAP for each specific IPD metanalysis. The DMP and SAP for each IPD meta-analysis will include: analysis objectives (primary, secondary); analytic sample inclusion and exclusion criteria; dependent and independent variable definitions; approaches to poorly or incongruently defined endpoints and other variables across studies; assessment of dependent and independent variable normality and functional transformation as required for advanced analysis; effect size (e.g., risk ratio, odds ratio); causal inference approaches, including model fitting, model diagnostics and approach to repeated measures; approach to missing data; evaluation of patterns of missingness; subgroup analyses; and evaluation of effect modifiers.

Note that objectives such as *in-vitro* assay development and evaluation may be performed off relatively small number of biospecimens (in contrast to objectives which seek to evaluate sensitivity, specificity, likelihood ratios of established assays).

### Generation of additional laboratory data from the specimen repository

While no new enrollment or participant data collection occurs in this repository protocol, secondary laboratory data, aligned with the secondary objectives listed, may be generated by current and future laboratory science collaborators using the specimens reposed in the ARI repository protocol.

## Discussion

This ARI Repository Protocol is expected to directly support further pandemic research response protocols, as well as follow-on lab-based studies (e.g., mechanistic studies for optimal vaccine response). The intended uses for this joint repository protocol include rapid assay development for emerging acute respiratory epidemic and pandemic pathogens. An illustration of the importance of having ‘on-demand’ biospecimen access was the use of the IDCRP ARIC protocol specimens [14] to rapidly support a high-throughput SARS-CoV-2 assay developed by USUHS investigators. This assay was then used across multiple IDCRP and non-IDCRP COVID-19 protocols to support multiple high tier publications and further diagnostic assay development [26, 45, 46].

In addition, this ARI Repository Protocol will allow rapid sample size calculations to inform new studies in MHS populations (for example, an interventional clinical trial or a diagnostic assay validation study), comparator population analyses (for example comparing the morbidity of a new pandemic respiratory virus against COVID-19 or seasonal influenza), and cross-validation of prognostic biomarkers. Furthermore, this repository protocol will permit IPD meta-analyses to answer ARI questions with greater statistical power than individual studies can permit alone. Such IPD meta-analyses are typically more epidemiologically valid than aggregate meta-analyses using study summary effect sizes [44]. These IPD analyses will be responsive to new respiratory virus epidemics and/or evolving MHS beneficiary health needs.

A strength of this ARI Repository Protocol is that many variables across the contributing protocols were measured in similar ways; for example, several studies used commonly worded CRF or survey response fields (e.g., FLUPRO scores across influenza and SARS-CoV-2 protocols [25, 31, 47]) and assays (e.g., common serology panels measured in the same USU lab across multiple COVID-19 protocols [26, 39, 42, 45, 46, 48].

The scientific objectives for this ARI Repository Protocol have clear military relevance to diseases of significant ADSM impact. The publication history of the contributing individual IDCRP protocols underscores the military relevance of this joint ARI Repository Protocol effort. In addition, this ARI Repository Protocol will help support future ARI pandemic research response. New laboratories interested in partnering on ARI investigations are welcome to inquire about collaborative opportunities aligned with the scientific aims of this biorepository.

## Table captions

**Table 1.**IDCRP Acute Respiratory Infection Study Characteristics

**Table 2.**IDCRP Acute Respiratory Infection Study Participant Information

**Table 3.**Protocol Objectives

## Data Availability

Data for the IDCRP-142 protocol study would be available from the Infectious Disease Clinical Research Program (IDCRP), headquartered at the Uniformed Services University of the Health Sciences (USUHS), Department of Preventive Medicine and Biostatistics. Review by the USU Institutional Review Board is required for use of the data collected. Furthermore, the clinical data set in some protocols which contribute to the IDCRP-142 protocol includes Military Health System data collected under Data Use Agreements that requires accounting for uses of the data. Clinical data requests may be sent to: Address: 6270A Rockledge Drive, Suite 250, Bethesda, MD 20817. Biospecimen requests by other institutions would be subject to specimen availability, protocol modification approvals, and material transfer agreements.

## List of abbreviations

ADSM: Active duty service member
ARI: Acute respiratory infection
ARIC: Acute Respiratory Infection Consortium
CAMP NYC: COVID-19 Antibody Prevalence in Military Personnel Deployed to New York
COVID-19: coronavirus disease 2019
CRF: Case report form
DCC: Data Coordinating Center
DMP: Data Management Plan
DoD: U.S. Department of Defense
DODM: U.S. Department of Defense manual
EPICC: Epidemiology, Immunology and Clinical Characteristics of Emerging Infectious Diseases with Pandemic Potential
FLUPRO: Influenza Patient-Reported Outcome
HIV: human immunodeficiency virus
HJF: Henry M. Jackson Foundation for the Advancement of Military Medicine
IDCRP: Infectious Diseases Clinical Research Program
IPD: individual participant-level data
IRB: Institutional Review Board
MHS: Military Health System
PAIVED: A Pragmatic Assessment of Influenza Vaccine Effectiveness in the Department of Defense
PASS: Prospective Assessment of SARS-CoV-2 Seroconversion
PBMC: Peripheral blood mononuclear cell
PI: Principal investigator
PISCES: Prospective Investigation of SARS-CoV-2/COVID-19 Epidemiology and Serology QA Quality Assurance
QC: Quality Control
QM: Quality Management
QMP: Quality Management Plan
SAP: Statistical analysis plan
SARS-CoV-2: Severe acute respiratory syndrome coronavirus 2
TOSCANA: The Observational Seroepidemiologic Study of COVID-19 at the United States Naval Academy
U.S.C.: United States Code
USNS: United States Navy Ship
U.S.S.: United States Ship
USU: Uniformed Services University
USUHS: Uniformed Services University of the Health Sciences

## Declarations

### Ethics approval and consent to participate

Provisions related to future use of data and specimens for research purposes were included in the Informed Consent Documents and text of the original protocols being merged for this repository (with the stipulation that the future use would be under a protocol reviewed by the USUHS IRB). This ARI Repository Protocol (IDCRP-142) was approved by USU Institutional Review Board and originating protocols have had modifications which refer to the movement of data and specimens into IDCRP-142.

## Authors’ contributions

Conceived and designed the study / experiments: SDP

Acquired data / performed experiments: RC, TL, AM, AF, EM, EDL, KP, EG, KM, MS, BA, DT, ROC, THB, Created detailed analysis plan and/or analyzed the data: SR, SDP

Interpreted findings: N/A

Contributed resources; reagents/materials/specimens: BB, EP, SM, OO, MG, MF Composed first draft of manuscript: SDP

Provided critical revisions and edits (scientific content) to provisional drafts: SDP

Reviewed and approved final version for submission RC, TL, AM, AF, EM, EDL, KP, EG, KM, MS, BA, DT, ROC, THB, MF

## Disclaimer

Views expressed are those of the authors and do not reflect the official policy of the Uniformed Services University of the Health Sciences, the Department of the Army, the Department of the Navy, the Department of the Air Force, the Department of Defense, the Defense Health Agency, or the U.S. Government and the Henry M. Jackson Foundation for the Advancement of Military Medicine, Inc. (HJF). The investigators have adhered to the policies for the protection of human subjects as prescribed in 45 CFR 46.

E.D.L, D.R.T, M.P.S, A.M, A.F, K.P, E.C.G, K.M, R.O., T.H.B, and E.M are U.S. Government employees or military service members. This work was prepared as part of the author(s) official duties. Title 17 U.S.C. § 105 provides that ‘Copyright protection under this title is not available for any work of the United States Government.’ Title 17 U.S.C. §101 defines U.S. Government work as work prepared by a military service member or employee of the U.S. Government as part of that person’s official duties.

