## Appendix A_Table 1 for "The Infectious Diseases Clinical Research Program Acute Respiratory Infection Repository Protocol: Opportunities to Understand Current and Future Epidemics"

**Table 1. IDCRP Acute Respiratory Infection Study Characteristics (up to n = 27, 231 total participants)**

| **Study (Protocol #)** | **Participant *n*** | **Pathogen(s)/Syndromes** | **Endpoints (data sources)** | **Specimen types (and associated lab data)** |
|---|---|---|---|---|
| ARIC (IDCRP-045) | 2,312 | Influenza, entero/rhinovirus, hCoV, RSV, hPIV, hMPV, ILI | Infection, severity (CRF) | Swabs (multiplex PCR, sequence data)[a] |
| H1N1 Flu Vaccine (IDCRP-053) | 132 | Influenza | Infection, immunity (CRF, MDR) | Sera (ELISA, CD4/HIV RNA levels), urine |
| EPICC (IDCRP-085) | 7,911 | COVID-19, CLI, other respiratory pathogens including entero/rhinovirus, influenza, hCoV, RSV, hPIV, hMPV | Infection, symptoms, acute/chronic complications/severity, immunity, linked transmissions (CRF/questionnaire, MDR, prospective imaging) | Sera (IgG binding/neutralizing antibodies), plasma (proteomics), PBMC (immunoseq, CITE-seq, functional assays), swabs (viral load, sequence data)[a], viral isolates (viral load, phenotype), PAXgene (RNAseq, host genome data)b, clinical residual specimens (viral load)[b] |
| PAIVED (IDCRP-120) | 15,449 | Influenza, COVID-19, CLI, ILI, entero/rhinovirus, hCoV, RSV, hPIV, hMPV | Infection (randomized trial primary endpoint), symptoms, immunity (CRF/questionnaire, | Sera (IgG binding/neutralizing antibodies), PBMC (immunoseq, functional assays), Swabs (viral load, sequence data)[a], host DNA from cheek swab, saliva (IgG, IgA) |
| CAMP_ NYC (IDCRP-125) | 372 | COVID-19 | Infection | Sera (IgG data only, no specimens) |
| PASS (IDCRP-126) | 280 | COVID-19, CLI | Infection, symptoms, acute severity, immunity (CRF/questionnaire) | Sera (IgG binding/neutralizing antibodies), PBMC (immunoseq, functional assays), saliva (IgG, IgA), clinical residual swabs |
| COMFORT (IDCRP-128) | 450 | COVID-19, CLI | Infection (CRF/questionnaire, MDR) | Sera (IgG) |
| TOSCANA (IDCRP-129) | 196 | COVID-19, CLI | Infection, immunity (CRF/questionnaire, MDR) | Sera (IgG), saliva (IgG, IgA) |
| PISCES (IDCRP-130) | 129 | COVID-19, CLI, influenza, entero/rhinovirus, hCoV, RSV, hPIV, hMPV hCoV, RSV, HPIV HMPV | Infection, immunity, linked transmissions (CRF/questionnaire) | Sera (IgG), swabs[a], saliva |

CLI = COVID-19-like illness; ILI = influenza-like illness, hCoV = non-SARS-CoV-2 endemic coronarviruses, RSV = respiratory syncytial virus, hPIV = parainfluenza viruses, hMPV = metapneumoviruses; PBMC = peripheral blood mononuclear cells; CRF = case report form; IgG = immunologobulin; MDR = Military Health System Data Repository

a. Swabs predominantly refer to upper respiratory swab specimens; b. Clinical residual specimens include upper respiratory swabs, bronchoalveolar lavage, stool and CSF
