## Appendix B_Table 2 for "The Infectious Diseases Clinical Research Program Acute Respiratory Infection Repository Protocol: Opportunities to Understand Current and Future Epidemics"

**Table 2. IDCRP Acute Respiratory Infection Study Participant Information (up to n = 27,231 total participants)**

| Study (Protocol #) | Age Range | Gender | Special Categories | Inclusion Criteria | Exclusion Criteria |
|---|---|---|---|---|---|
| **ARIC (IDCRP-045)** | **17-year-old military trainees 18-65** | **Male/ Female** | **** Resident/ Trainee<br>** Active-Duty Military Personnel** | **Inclusion Criteria:**<br>1. Between 18-65 years of age (inclusive), or a 17-year-old military service member.<br>2. Have an acute influenza -like illness (ILI) or SARI symptomatic for </= 7 days as defined in appendix A.<br>3. Eligible for care in DOD facilities (DEERS eligible). | No exclusion criteria has been added to this protocol. |
| **H1N1 Vaccine (IDCRP-053)** | **18-50** | **Male/ Female** | **** Military Beneficiary** | **Inclusion Criteria:**<br>1. 18-50 Years of age<br>2. Receiving the novel H1N1 vaccine (killed formulation)<br>as part of routine clinical care.<br>3. A military beneficiary who expects to remain in the local<br>area for the next 6 months. | **Exclusion Criteria:**<br>1. Are a healthcare worker who sees patients (Healthcare workers may be particularly likely to be exposed to influenza as part of their work and have altered immune responses due to natural exposures)<br>2. Have had an acute febrile illness within 30 days prior to HlNl vaccination (e.g., pneumonia, influenza)<br>3. Have any of the following medical conditions:<br>** Diabetes type 1or type 2<br>** Systemic steroid or immunosuppressive medication use within past 4 weeks<br>** Active diagnoses of a cancer (non-melanoma skin cancer allowed).<br>** History of organ transplant<br>** Chronic active hepatitis B or C<br>4/ Actively use illicit drug use or abuse alcohol<br>5. Have had a blood transfusion within last year<br>6. Are allergic to eggs<br>7. Have had a previous significant adverse reaction to the influenza vaccination |

| | | | | | |
|---|---|---|---|---|---|
| **EPICC (IDCRP-085)** | **0-75+** | **Male/ Female** | **** Minors/ Children<br>** Active-Duty Military Personnel<br>** Persons with Impaired Decisional Capacity<br>** Pregnant Women, Fetuses, and Neonates** | **Inclusion Criteria:**<br>MHS beneficiaries of any age who meet at least one of the following criteria:<br>1. Laboratory-confirmed presence of the pathogen of interest<br>2. Meet criteria for testing for the pathogen of interest, as identified per current CDC guidelines.<br>3. Received vaccine for the pathogen of interest.<br>**Additional Inclusion criteria for Online Enrollment:**<br>1. Able to receive email communications and respond to web-based questio0nnaires.<br>2. Greater than or equal to 18 years of age. | **Exclusion Criteria:**<br>1. Individuals who decline participation in the study.<br>2. Individuals who the study investigators believe ar |
| **PAIVED (IDCRP-120)** | **18-75+** | **Male/ Female** | ****Students<br>** Resident/ Trainee<br>** Cadets/ Midshipmen<br>** Active-Duty Military Personnel** | **Inclusion Criteria:**<br>1. Eligible for care in DOD Facilities (DEERS eligible)<br>2. Greater than or equal to 18 years of age<br>3. At a participating MTF site for the purpose of receiving a seasonal influenza vaccination<br>4. Able to speak English and able to provide informed consent.<br>5. Able to receive and respond to texts and/or emails, or a military recruit. | **Exclusion Criteria:**<br>1. Adults intending to receive or who have received<br>2. Adults who have already received a Flu vaccine within the current season.<br>3. Individual who cannot receive a flu vaccine or standard dosing due to another medical condition.<br>4. Allergic to gentamicin, polymyxin, and/or neomycin.<br>5. Individuals who fail to meet the inclusion criteria. |
| **CAMP_NYC (IDCRP-125)** | **18-75+** | **Male/Female/ Other** | **** Active-Duty Military Personnel** | **Inclusion Criteria:**<br>COHORT 1: All Active Duty Army personnel deployed to NYC from Fort Sam Houston (San Antonio, TX), Fort Campbell, KY, and Fort Hood, TX.<br>COHORT 2: Individuals admitted to the Javits Convention Center with nasal swab PCR proven COVID-19 infection. | **Exclusion Criteria:**<br>1. COHORT 1: Personnel deployed from areas with high rates of community spread of COVID-19 at the time of their arrival to NYC.<br>2. COHORT 2: Patients without a confirmed COVID-19 positive test<br>3. COHORT 2: Symptom onset < 4 days prior<br>4. COHORT 2: Patients who are not fluent in English<br>5. COHORT 2: Altered mental status, dementia, or impaired cognition (as assessed by the study investigator or indicated in their medical records)<br>6. COHORT 2: Age less than 18 (also makes them ineligible for admission to the Javits Convention Center. |

| | | | | | |
|---|---|---|---|---|---|
| **PASS (IDCRP-126)** | **18-75+** | **Male/ Female** | **** Employees-Civilian** <br> **** Employees-Contractor** <br> **** Resident/ Trainee** <br> **** Active-Duty Military Personnel** | **Inclusion Criteria:** <br> 1. Age, greater than or equal to 18 years old. <br> 2. Generally healthy <br> 3. Health care providers (nurses, doctors, physician assistants, respiratory therapists, occupational therapists, and medical technicians) at WRNMMC. <br> 4. Willingness and ability to return for scheduled follow-up visits, with assurance of follow-up at least for the first three months. | **Exclusion Criteria:** <br> 1. Immunocompromised state or immune modulating medications - presence of a disease that is actively causing severe immune suppression 20 mg prednisone or greater daily for over one month, chemotherapy, cytokine inhibitors, or agents that reduce T cell or B cell numbers or function. <br> 2. Symptoms of fever, cough, anorexia, myalgias, chills, shortness of breath, anosmia, sore throat, rhinorrhea, or diarrhea at enrollment. <br> 3. Presence of fever (T > 100.4 °F) on screening vital signs. <br> 4. Known prior diagnosis with COVID-19. <br> 5. Positive SARS-CoV-2 IgG by multiplex coronavirus serology assay. <br> 6. Participation in a COVID-19 vaccine or prophylactic antibody study. |
| **COMFORT (IDCRP-128)** | **18-75** | **Male/ Female** | **** Active-Duty Military Personnel** | **Inclusion Criteria:** <br> 1. Active-Duty US Navy Active-Duty and Reserve personnel participating in the COVID-19 related missions on the USNS COMFORT and MERCY hospital ship. <br> 2. Willing to provide informed consent and comply with study procedures (questionnaire and blood draw) | No exclusion criteria has been added to this protocol. |
| **TOSCANA (IDCRP-129)** | **17-27** | **Male/ Female** | **** Cadets/ Midshipmen** | **Inclusion Criteria:** <br> 1. US Naval Academy Midshipman, male or female subject, 17-27 years of age, inclusive, at the time of screening. <br> 2. Able to view video link advertising the study and to read and complete on-line informed consent document. <br> 3. Informed of the nature of the study and has agreed to and is able to read, review, and sign the informed consent document prior to screening. <br> 4. Free of known significant health problems as established by the requirements to be enrolled in a military service academy before entering into the study | **Exclusion Criteria:** <br> 1. Not able to view video link advertising the study and to read and complete on-line informed consent document. <br> 2. Individuals who fail to meet the inclusion criteria. |

| | | | | | |
|---|---|---|---|---|---|
| **PISCES (IDCRP-130)** | **Children:**<br>**>2-17**<br>**Adults:**<br>**18-75+** | **Male/ Female** | **** Minors/ Children**<br>**** Students**<br>**** Employees-Civilian**<br>**** Employees-Contractor**<br>**** Resident/ Trainee**<br>**** Active-Duty Military Personnel**<br>**** Pregnant Women, Fetuses, and Neonates** | **Inclusion Criteria:**<br>1. USU affiliate*, male or female18 years of age (primary enrollee),>*USU affiliate: A USU faculty, staff, student, Active-Duty military, or contractor with the USU Bethesda, MD campus as their primary place of duty outside the home and with a current functioning USUHS.edu email address.<br>2. Household member of the primary enrollee, >24 months of age, parent or guardian of the individual is 18 years of age.<br>3. Volunteer or parent/legal guardian able to view a video link advertising the study and to read and complete on-line informed consent document.<br>4. Volunteer or parent/guardian informed of the nature of the study and has agreed to and is able to read, review, and sign the informed consent document prior to screening. | **Exclusion Criteria:**<br>1. Volunteer or parent/legal guardian not able to view video link advertising the study and to read and complete on-line informed consent document.<br>2. Current participation in COVID-19 vaccine trials or any prospective studies that includes investigational pre- or post- exposure prophylaxsis products. Future vaccine receipt or receipt of any prophylaxis product will not be grounds for removal from the study.<br>3. < 24 month of age household member |
