## Appendix C_Table 3 for "The Infectious Diseases Clinical Research Program Acute Respiratory Infection Repository Protocol: Opportunities to Understand Current and Future Epidemics"

**Appendix C. Table 3, Protocol Objectives**

| Protocol Objectives/ Aims |
|---|
| **EPICC (IDCRP-085) Epidemiology, Immunology and Clinical Characteristics of Emerging Infectious Diseases with Pandemic Potential** |
| **For COVID-19 / SARS-CoV-2 infection, clinical and demographic data will be evaluated, along with laboratory findings as appropriate, to assess key endpoints of interest to include, but not limited to, the following:**<br>Incubation period<br>Duration of illness / symptoms<br>Duration of functional disability<br>Duration of shedding from respiratory and gastrointestinal tract<br>Incidence and duration of viremia<br>Predictors of severe disease: clinical signs, symptoms, laboratory features; host factors, eg smoking, vaping, alcohol consumption, comorbid illness/conditions, occupational health factors influencing lung disease; host biomarkers; virologic (strain/virulence) attributes<br>Supportive management required: O2, HFNC, NIPPV, ventilation; hemodynamics Complications: e.g., bacterial pneumonia; bacteremia; AKI; liver injury; CNS involvement, etc.<br>Outcomes: resolution of symptoms; return to pre-illness functional status; requirement<br>for O2; development / exacerbation of reactive airway disease; pulmonary function; resolution of medical complications; new Rx requirements, eg bronchodilators; other sequelae<br>Development of immunity (i.e., correlates), kinetics, duration of antibody detection<br>/protection including in response to Covid vaccine; evaluation of reinfection cases/risk; estimation of vaccine effectiveness<br>Relationship between acute illness (or pre-illness) HCoV titer and infection, outcome, virologic parameters<br>Costs of care, costs of duty days lost |
| **PAIVED (IDCRP-120) A Pragmatic Assessment of Influenza Vaccine Effectiveness in the DoD** |
| **Specific Aim #1**: Comparison of the relative effectiveness (prevention of laboratory-confirmed influenza illness) of three types of licensed seasonal influenza vaccines.<br>**Specific Aim #2**: (Immunogenicity substudy) Determine whether cell-culture-based, egg-based and recombinant influenza vaccines give comparable HI and PVN titers to egg- and cell-matched vaccine antigens. An outcome of this objective is the potential to determine whether cell-culture-based vaccine antigens can provide broader coverage of circulating viruses than egg-based vaccine antigens.<br>**Specific Aim #3**: To determine if the impact of the influenza vaccine on disease burden and the attributable healthcare costs differs by product type.<br>**Specific Aim #4***:* To evaluate the association host single nucleotide polymorphisms (SNPs) and immune responses to the influenza vaccine, influenza acquisition and influenza severity.<br>**Specific Aim #5**: Assess the burden of COVID-19 and explore the inter-relationship between influenza and COVID-19. |
| **PASS (IDCRP-126) Prospective Assessment of SARS-CoV-2 Seroconversion** |
| **Specific Aim #1**: Determine the frequency of asymptomatic and pauci-symptomatic SARS-CoV-2 infection in a cohort of healthcare workers.<br>**Specific Aim# 2**: Investigate the serological footprint of anti-S glycoprotein antibodies to HCoV-OC43, -HKU1, -229E, and -NL63, and determine whether pre-existing antibodies to these HCoVs are associated with altered COVID-19 disease course.<br>**Specific Aim #3**: Determine if pre-existing immune responses to seasonal coronaviruses affect magnitude and duration of antibody and T-cell responses to SARS-CoV-2 vaccination. |
| **ARIC (IDCRP-045) The Acute Respiratory Infection Consortium** |

| |
|---|
| **Primary Objective:** Develop a consortium of DoD research sites, which are capable of collecting detailed prospective clinical data and biologic samples from subjects with respiratory inf ections, particularly ILI, for analysis of the impact of these illnesses on our active-duty members and their families.<br>Secondary Objective A: Develop and validate an influenza severity scale. A preliminary severity scale for ILI will be developed and applied. The scale will be based on subjective and objective findings, which are easily measured and duplicated (as many numeric scales as is practical).<br>Secondary Objective B: Study the relationship between health and fitness of young adults to outcomes of influenza and other respiratory pathogens. Secondary Objective C: Examine the relationship of cell-mediated and humoral immune response to the severity of influenza and other respiratory pathogens.<br>Secondary Objective D: Describe patterns of viral shedding in different influenza types and subtypes. Specifically, describe the frequency of asymptomatic shedding, identify possible transmission associated with asymptomatic shedding, and describe the duration of shedding in subjects with different influenza viruses and across a wide spectrum of clinical disease.<br>Secondary Objective E: Correlate clinical severity and cytokine/humoral response with underlying host genotype.<br>Secondary Objective F: To determine the representativeness of the cohort to the referent population with regards to demographics, ILI/SARI severity, and pathogen distribution.<br>Secondary Objective G: To investigate interaction between influenza and other respiratory pathogens.<br>Secondary Objective H: To describe the pathogens associated with cases requiring intensive care or mechanical ventilation.<br>Secondary Objective I: To study the use of antimicrobial agents in the treatment of ILI and SARI.<br>Secondary Objective J: To study inflammatory immune responses to pathogens associated with SARI.<br>Secondary Objective K: To characterize antimicrobial sensitivity patterns of ILI and SARI pathogens.<br>Secondary Objective L: To estimate influenza vaccine and adenovirus vaccine effectiveness.<br>Secondary Objective M: To explore ILI- and SARI-associated health care utilization and economic burden, duty and work absences associated with ILI and SARI.<br>Secondary Objective N: To examine site of care decisions (inpatient vs. outpatient) related to SARI and ILI. |
| **COMFORT (IDCRP-128) Seroprevalence of novel coronavirus antibodies among personnel deployed on the USNS Comfort and Mercy during the COVID 19 pandemic** |
| **Primary Objective:** Determine the seroprevalence of COVID-19 antibodies among US Navy personnel deployed to Los Angeles or New York City on the USNS MERCY and COMFORT hospital ships in support of COVID-19 outbreaks in those cities.<br>**Secondary objectives:** Compare the seroprevalence of COVID-19 antibodies between the Navy personnel deployed on the USNS MERCY and those on the COMFORT. Correlate the likelihood of COVID-19 antibody prevalence with assigned duties (nonclinical, medic, nursing, physician), workspace, social circles, berthing, and known exposure to COVID-19 infected patients. Determine the incidence of medically-attended, laboratory-confirmed COVID-19 among enrolled personnel using medical record abstraction 1-year post-enrollment. Compare post-deployment seroprevalence to a convenience sample of those crew members with available DoD serum repository samples within 1-2 months prior to deployment. |
| **PISCES (IDCRP-130) Prospective Investigation of SARS-CoV-2 / COVID-19 Epidemiology and Serology** |
| **Specific Aims:** To determine the prevalence and incidence of SARS-CoV-2 infection. To evaluate the association between baseline SARS-CoV-2 antibody seroprevalence and attack rate of SARS-CoV-2 over the course of a respiratory disease season. To describe the association between occupational-, household-, and community-level risk factors and the risk of SARS-CoV-2 infection. To describe the functional impact of SARS-CoV-2 and non-SARS-CoV-2 respiratory pathogens on the health of the USU community across one respiratory disease season. To characterize the elicitation and durability of the host immune response to SARS-CoV-2 and the influence of symptom/disease severity on the magnitude of the response. To examine the correlations between systemic and mucosal immune response following exposure to infection with SARS-CoV-2. |
| **TOSCANA (IDCRP-129) The Observational Seroepidemiologic Study of COVID-19 at the United States Naval Academy** |

| |
|---|
| **Specific Aims**: To determine the prevalence and incidence of SARS-CoV-2 infection among students at a military service academy. To evaluate the association between baseline SARS-CoV-2 antibody seroprevalence and attack rate of SARS-CoV-2. To evaluate the association between baseline SARS-CoV-2 antibody seroprevalence and rate of medically-attended ARI due to SARS-CoV-2. To describe the proportion and temporal distribution of medically-attended ARI due to SARS-CoV-2 relative to that of common seasonal respiratory pathogens. To describe individual-level risk factors (i.e. host genetic determinants) associated with progression to clinically-significant COVID-19. To evaluate COVID-19 mitigation approaches and the relative impact of these strategies on rates of all-cause as well as SARS-CoV-2 medically-attended ARI in a university dormitory setting. |
| **H1N1 Vaccine (IDCRP-053) Immunogenicity of Novel H1N1 Vaccination among HIV-Infected Compared to HIV-Uninfected Persons** |
| **Primary Objective:** To compare the immunogenicity via anti-hemagglutinin responses following H1N1 vaccination between HIV positive and negative persons. Null Hypothesis (Ho): The immunogenicity of the H1N1 vaccine by anti-hemagglutinin seroresponse between these groups will not be significantly different.<br>**Secondary Objectives:**<br>1. To compare the immunogenicity via HAI titer levels, microneutralization seroresponses and titer levels, and cellular responses following H1N1 vaccination (i.e., visit 1 to visit 2 changes) between HIV positive and negative persons. Null Hypothesis (Ho): The immunogenicity of the H1N1 vaccine by HAI titer levels, microneutralization seroresponse and cellular responses between these groups will not be significantly different.<br>2. Among those undergoing vaccination with the seasonal influenza vaccine during the current influenza season, to compare the presence of a positive seroresponse between HIV positive and negative persons. Ho: The presence of a seroresponse to seasonal influenza vaccination between these groups will not be significantly different.<br>3. To evaluate the effect of pre-existing anti-influenza immunity and recent history of seasonal influenza vaccination on seroresponses to the H1N1 influenza vaccine among both HIV positive and negative persons. Ho: Prior anti-influenza immunity or recent seasonal influenza vaccination will have no impact on H1N1 vaccine immune responses.<br>4. To compare the durability of the H1N1 immunologic responses at 6 months post-vaccination between HIV-infected and uninfected persons. Ho: The durability of the immune responses to the H1N1 vaccine as measured by antibody seroresponses and cellular responses between these groups will not be significantly different.<br>5. To evaluate the number of ILIs and documented influenza cases among HIV-infected and uninfected persons after initial vaccination, and to genetically characterize the influenza strains causing ILI events in our study cohort. |
| **CAMP_NYC (IDCRP-125) COVID-19 Antibody Prevalence in Military Personnel Deployed to New York (Determination of novel coronavirus antibody prevalence among personnel in deployed in response to the New York City COVID-19 pandemic)** |
| **Primary Objective**: To determine the seroprevalence of COVID-19 antibodies among active-duty Army personnel deployed to New York City to respond to the COVID-19 pandemic prior to their redeployment.<br>**Secondary objectives:**<br>1. To correlate the likelihood of COVID-19 antibody prevalence with frequency of exposure to COVID-19 infected patients.<br>2. To correlate the likelihood of COVID-19 antibody prevalence with type of duties (nonclinical, medic, nursing, physician).<br>3. To correlate the likelihood of COVID-19 antibody prevalence with history of performance aerosolizing procedures.<br>4. To correlate the presence COVID-19 antibody prevalence with history of COVID-like symptoms.<br>5. To evaluate the performance characteristics of Premier Biotechnology qualitative COVID-19 antibody test.<br>6. To determine the rate of viral RNA detection in patients with confirmed COVID-19 and correlate with the presence and titer of serum antibodies. |
